# Melting curve analysis reveals false-positive norovirus detection in a molecular syndromic panel

**DOI:** 10.1101/2024.02.23.24303102

**Authors:** Nancy Matic, Tanya Lawson, Matthew Young, Willson Jang, Jennifer Bilawka, Leah Gowland, Gordon Ritchie, Victor Leung, Michael Payne, Aleksandra Stefanovic, Marc G. Romney, Christopher F. Lowe

**Affiliations:** Division of Medical Microbiology and Virology, St. Paul’s Hospital, Vancouver, Canada; Department of Pathology and Laboratory Medicine, University British Columbia, Vancouver, Canada

**Keywords:** norovirus, syndromic, panel, molecular, false-positive

## Abstract

**Background:** Molecular syndromic panels can improve rapidity of results and ease clinical laboratory workflow, although caution has been raised for potential false-positive results. Upon implementation of a new panel for infectious diarrhea (BioFire® FilmArray® Gastrointestinal [GI] Panel, bioMérieux) in our clinical laboratory, a high number of stool samples with norovirus were detected.

**Objectives:** To investigate positive percent agreement and the false-positive rate of norovirus detected by the multiplex Biofire GI panel compared to a singleplex commercial assay.

**Study design:** From October 2023 to January 2024, all prospective stool samples with a positive norovirus result by BioFire had melting curves reviewed manually using the BioFire FilmArray Torch System. Stool samples further underwent testing by a supplementary real-time RT-PCR assay (Xpert® Norovirus, Cepheid) for comparative analysis.

**Results:** Of the 50 stool samples with norovirus detected by BioFire, 18 (36%) tested negative by Xpert (deemed “false-positives”). Melting curve analysis revealed nearly all of these samples had atypical melting curve morphologies for the “Noro-1” target on BioFire (16/18, 89%), which was statistically significant (Odds Ratio 173.2, 95% CI [22.2, 5326.9], *p*<0.0001). Stool samples with multiple pathogens including norovirus detected by BioFire were not more likely to produce false-positive norovirus results (Odds Ratio 1, 95% CI [0.3, 3.3], *p*=1).

**Conclusions:** Although not described in the manufacturer’s *Instructions for Use*, we propose routine manual review of melting curves for the BioFire GI panel prior to reporting, to avoid potential false-positive norovirus results.

## Background

The diagnosis of infectious gastroenteritis has evolved rapidly in the past decade from culture-based workup to molecular syndromic panels. The conversion to molecular testing for infectious gastroenteritis enables rapid turnaround time for results, increased sensitivity for pathogen detection, and optimize laboratory productivity[1,2]. However, depending on the patient’s risk factors and clinical presentation, the pre-test probability of each target on the panel varies, which may affect positive predictive value[3].

Norovirus, a non-enveloped RNA virus, is often included in molecular panels as a common cause of viral gastroenteritis[4]. Historically, within healthcare facilities, norovirus testing had been limited to situations where there was suspicion for patient-to-patient transmission or outbreak[5]. In British Columbia, there has been a recent shift to molecular panels as the primary diagnostic modality for infectious gastroenteritis[6]. As a result, patients presenting with acute gastrointestinal symptoms are now routinely tested for norovirus, in addition to other gastrointestinal pathogens. Although the community burden of disease is seasonal and increased norovirus detection would be expected between November to April[7], our clinical laboratory noted a significant increase in norovirus detection since the routine implementation of the molecular infectious diarrhea panel.

## Objectives

We investigated percent positive agreement and the false-positive rate of norovirus detected by the multiplex BioFire GI panel compared to a singleplex commercial assay for prospective stool samples.

## Study design

From October 4, 2023 to January 26, 2024, stool collected in sterile screw-top containers from patients presenting to two adult tertiary care hospitals with acute gastroenteritis were received in our clinical microbiology laboratory. Stool samples were rejected if they were not loose/watery (Bristol type 6 or 7) or collected >72 hours after admission. Neat stools were subsequently transferred into 2 mL of Cary-Blair media (FecalSwab™, Copan Diagnostics) and tested by the BioFire® FilmArray® Gastrointestinal (GI) Panel (bioMérieux) according to manufacturer’s instructions. Melting curves were reviewed on the BioFire® FilmArray® Torch System (version 3.2.4.0). All samples with a positive norovirus result by the BioFire panel were then tested by a supplementary norovirus real-time RT-PCR platform (Xpert® Norovirus, Cepheid) according to manufacturer’s instructions for comparative analysis. Statistical analysis was performed using RStudio (2022.2.1.461).

## Results

A total of 800 stool specimens were received in the laboratory during the study period; 97 samples met rejection criteria and were excluded from testing. The remaining 703 samples from 656 unique patients (Table 1) underwent testing by the BioFire panel; 7.1% (50/703) of samples had a positive norovirus result. Among these samples, 50% (25/50) had norovirus detected in combination with one or more additional pathogens (Table 2). The pathogens most frequently detected in combination with norovirus in our patient population were *Shigella*/Enteroinvasive *E. coli* (EIEC, 9, 36%), Enteroaggregative *E. coli* (EAEC, 7, 28%), and *Clostridioides difficile* (6, 24%).

**Table 1:** Demographics of the patient population presenting with acute gastroenteritis in this study (N=656)

| Parameter | No. (%) of samples |
| --- | --- |
| *Sex* |  |
| Male | 367 (55.9) |
| Female | 289 (44.1) |
| *Age (years)* |  |
| <5 | 0 |
| 5-17 | 6 (0.9) |
| 18-35 | 136 (20.7) |
| 36-55 | 209 (31.9) |
| 56-75 | 212 (32.3) |
| >75 | 93 (14.2) |
| *Specimen Collection Location* |  |
| Emergency department | 430 (65.5) |
| Outpatient clinic | 114 (17.4) |
| Inpatient ward | 112 (17.1) |

**Table 2:** Melting curve analysis of norovirus results by the BioFire® FilmArray® Gastrointestinal (GI) Panel and comparison to the Xpert® Norovirus assay.

| BioFire GI Results |  |  |  | Xpert Norovirus Results |  |  |
| --- | --- | --- | --- | --- | --- | --- |
| No. of targets detected | No. of samples | Melting curve analysis with atypical curve for “Noro-1” target |  | No. Detected | No. not Detected | % Detected |
|  |  | Yes | No |  |  |  |
| *Multiple (incl. Norovirus)* |  |  |  |  |  |  |
| 5 | 1 | 1 | 0 | 0 | 1 | 0 |
| 4 | 2 | 0 | 2 | 2 | 0 | 100 |
| 3 | 10 | 1 | 9 | 7 | 3 | 70.0 |
| 2 | 12 | 5 | 7 | 7 | 5 | 58.3 |
| *Single (only Norovirus)* |  |  |  |  |  |  |
| 1 | 25 | 10 | 15 | 16 | 9 | 64.0 |
| **Total** | **50** | **17** | **33** | **32** | **18** | **64.0** |

Of the 50 samples that underwent supplementary norovirus RT-PCR testing by the Xpert assay, 64% (32/50) confirmed positive by Xpert (Table 2). Cycle threshold (Ct) values on the Xpert ranged from 13 to 38 (median 22); genogroup I (GI) comprised 18.8% (6/32) of cases, genogroup II (GII) comprised 71.9% (23/32) of cases, and notably 9.4% (3/32) of cases were mixed GI and GII norovirus infection. The presence of multiple targets detected by BioFire did not significantly increase the likelihood of a negative Xpert result compared to samples with only a single norovirus target detected (Odds Ratio 1.0, 95% CI [0.3, 3.3], *p* 1.0). Review of BioFire melting curves demonstrated atypical curves of the “Noro-1” target (Figure 1) for nearly all samples that tested negative by Xpert (16/18, 88.9%) and for one sample that tested positive by Xpert (1/32, 3.1%). This association was statistically significant (Odds Ratio 173.2, 95% CI [22.2, 5326.9], *p* <0.0001).

**Figure 1:**
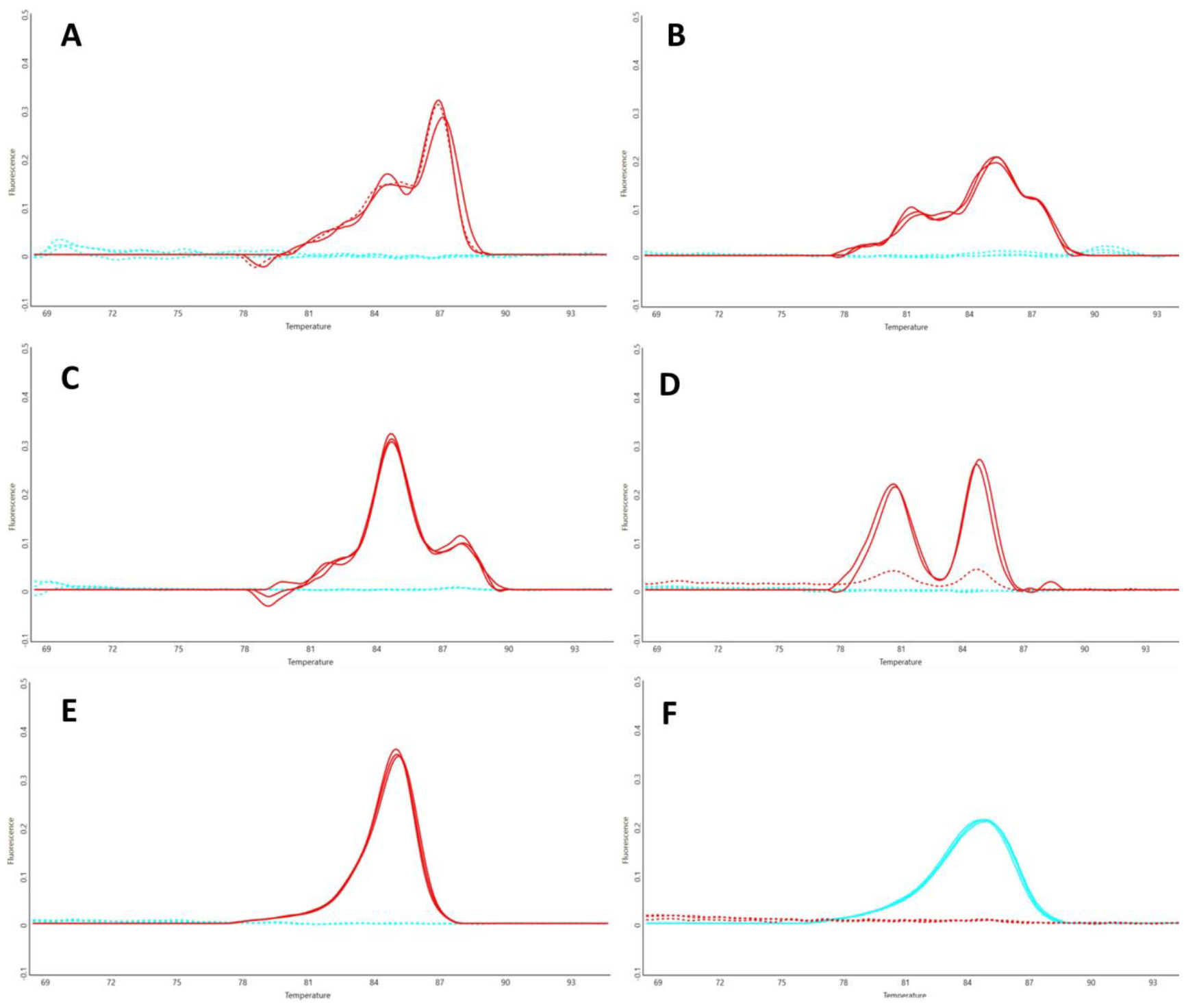
Examples of atypical, multi-modal melting curves for the “Noro-1” target (A, B, C, D) versus acceptable melting curves for the “Noro-1” (E) and “Noro-2” (F) targets on the BioFire® FilmArray® Gastrointestinal (GI) Panel. “Noro-1” melting curves are displayed in red, “Noro-2” melting curves are displayed in blue.

## Discussion

Molecular syndromic panels have remarkable clinical and operational advantages, including rapidity of results and ease of laboratory workflow; however, caution has been advised when launching new molecular syndromic panels, with concern for potential decreased sensitivity compared to singleplex assays and the inclusion of multiple targets which may increase risk for false-positive results[8,9]. Concerns about specificity of norovirus results on the BioFire GI panel have been reported previously[10,11], with false-positive results observed at a rate of 20-25%. In this study, we observed a similar though slightly higher false-positive rate of 36% in comparison to the Xpert Norovirus assay, a well-studied and widely-accepted real-time RT-PCR test[12,13]. The manufacturer has recently issued a correction notice regarding the risk for false-positive norovirus results with the BioFire GI panel[14], suggesting supplementary testing could be considered when results are not consistent with clinical presentation.

False-positive norovirus results were previously well-described in laboratory-developed end-point PCR assays, with nonspecific amplification of human DNA in stool samples[15]. This occurred most often in stool samples from patients with bacterial gastroenteritis, potentially due to the increased GI mucosal inflammation and leukocytes in these stool samples. Our study did not reveal an increased rate of false-positive norovirus results among samples with additional bacterial pathogens detected, suggesting that cross-reactivity likely did not occur with these particular pathogens; however, it is possible cross-reactivity occurred with other organisms or commensals in the stool, or other unknown factors.

Melting curve analysis was a striking finding in our study revealing the majority of samples with false-positive norovirus results had atypical multi-modal “Noro-1” melting curves on the BioFire panel compared to true-positives with the expected single peak. The atypical appearance suggests nonspecific incidental amplification, but has not been previously well-described for the BioFire panel. It should be noted that one sample in this study demonstrated an atypical melting curve for the “Noro-1” target only, but did test positive for norovirus GI by Xpert with a late Ct value of 36. The clinical relevance of this result is questionable, as low viral loads in stool may be consistent with asymptomatic viral shedding up to 55 days post-infection[16] and are less associated with symptomatic disease[17]. Additionally, two samples in this study had acceptable single-peak BioFire melting curves for the “Noro-2” target, yet tested negative by Xpert. Each of these were positive for additional BioFire panel targets (one with *Shigella*/EIEC and Enteropathogenic *E. coli* [EPEC], the other with EAEC and Enterotoxigenic *E. coli* [ETEC]). It is possible these samples had low amounts of norovirus near the limit of detection of the Xpert assay, and the patients’ symptoms may be more accurately attributed to bacterial infections.

This study has several limitations, including limited sample size due to the nature of the prospective testing since the BioFire GI panel was implemented in October 2023; however, testing prospective samples in parallel allowed us to avoid multiple freeze/thaw cycles of retrospective samples which may impact analytical results. We did not test all prospective stool samples by the Xpert Norovirus test; therefore, this study does not evaluate positive and negative predictive values of the BioFire GI Panel, but rather focuses on the investigation of potential false-positive norovirus results. A limitation of BioFire technology is the inability to access amplification data (e.g., cycle threshold values or amplification curves) to further evaluate the reliability of results. Lastly, this study did not include melting curve analysis for other targets on the BioFire GI Panel.

Molecular syndromic panels have great potential to enhance diagnostics and patient care, with appropriate laboratory stewardship and interpretation of results. The ease of use of the BioFire FilmArray Torch System allows for immediate visualization of melting curves before reporting. Although not described in the manufacturer’s *Instructions for Use*, we propose routine review of melting curves for the BioFire GI panel prior to reporting, to avoid potential false-positive norovirus results.

## Data Availability

All data produced in the present work are contained in the manuscript.

